# Estimating weight in patients of all-ages: a prospective study in Rwanda

**DOI:** 10.1101/2020.02.29.20029371

**Authors:** Giles N Cattermole, Appolinaire Manirafasha

**Author notes:** **Corresponding author** Giles N Cattermole, Emergency Department, Princess Royal University Hospital, Orpington BR6 8ND, UK.

## Abstract

**Introduction:** Weight estimation of both adult and paediatric patients is often necessary in emergency or low-resource settings when it is not possible to weigh the patient. There are many methods for paediatric weight estimation, but no standard methods for adults. PAWPER and Mercy tapes are used in children, but have not been assessed in adults. The primary aim of this study was to assess weight estimation methods in patients of all ages.

**Methods:** Patients were prospectively recruited from emergency and out-patient departments. Subjects (or guardians) were asked to estimate weight. Investigators collected weight, height, mid-arm circumference (MAC) and humeral-length data. In all subjects, estimates of weight were calculated from height and MAC (PAWPER tapes), MAC and humeral-length (Mercy tape). In children, Broselow tape and age-based formulae were also used. Primary outcome measures were proportions of estimates within 10%, 20% and 30% of actual weight (p10, p20, p30).

**Results:** 947 subjects were recruited: 307 children, 309 adolescents and 331 adults. For p20, the best methods were: in children, guardian estimate (90.2%) and PAWPER XL-MAC (89.3%); in adolescents, PAWPER XL-MAC (91.3%) and guardian estimate (90.9%); in adults, subject estimate (98.5%) and PAWPER XL-MAC (83.7%).

**Conclusion:** This is the first prospective study of weight estimation methods in Rwanda, and the first adult study of PAWPER and Mercy tapes. In children, age-based rules performed poorly. In patients of all ages, the PAWPER XL-MAC and guardian/subject estimates of weight were the most reliable and we would recommend their use in this setting.

## INTRODUCTION

Paediatric weight is often estimated in emergency situations using methods based on age (eg, the APLS formulae[1]) or height (eg, the Broselow tape[2]) when direct measurement is not possible. Methods based on mid-arm circumference (MAC) have also been developed.[3, 4] The Mercy [4] and PAWPER [5] tapes utilise both length-based and habitus-based body measurements in order to estimate weight. The Mercy tape uses humeral length and MAC; the PAWPER tape uses height and an estimate of overall body habitus. The PAWPER tape has recently been adapted to use MAC instead of the habitus estimate.[6] A retrospective study based on case notes of paediatric patients in Rwanda derived a new age-based formula for children aged 1-10 years old.[7]

Many drug and fluid regimes are weight-dependent in adults too. In our setting, the most frequently prescribed weight-dependent drugs are for rapid sequence intubation or procedural sedation, where it is especially important to ensure we give the appropriate doses. However, there are no accepted standard adult weight-estimation tools. Age-based methods are not appropriate, and the Broselow tape has been shown to be inappropriate for people over 10 years old, because the large majority are too tall to fit the tape.[8] A MAC-based adult weight estimation formula has been derived and validated using data from the American NHANES (National Health and Nutrition Examination Survey) database, [9] and a height-based formula for adult weight estimation has recently been derived in Nigeria [10].

In resource limited settings, weight estimation methods for patients of all ages are often helpful even in non-emergency situations, because accurate weighing facilities may not be readily available.[6]

Neither the PAWPER nor Mercy tapes have yet been studied in an adult population. To date in Rwanda, there have been no weight estimation studies in adults or adolescents, no prospective studies in children, and no studies assessing the MAC formula, Mercy or PAWPER tapes in children.

The aims of this study were:

1. To compare existing methods of weight estimation prospectively in children, adolescents and adults in Rwanda.
2. To validate the use of the Mercy and PAWPER tapes in adults and adolescents.
3. To validate the retrospectively derived Rwanda age-based rule in children.

## METHODS

This was a prospective observational study performed in Rwanda’s public tertiary referral hospital, Centre Hospitalier Universitaire de Kigali (CHUK). Data were collected over a three month period from December 2016 to February 2017.

Two investigators recruited a convenience sample of patients from the waiting rooms of the adult and paediatric emergency and out-patient departments. Patients were recruited in three groups, aiming for an approximately even spread of ages within each of three categories: children (aged 1-9 years last birthday), adolescents (10-15 years last birthday) and adults (16 years old and above). Pregnant women, patients in extremis, and infants were excluded.

Guardians provided an estimate of weight for subjects under 16 years old; adults provided an estimate of their own weight. Investigators used the graphical descriptive scale of body habitus on the PAWPER tape, to allocate children and adolescents to one of seven body habitus categories.[11] Subjects were then weighed to 0.1 kg using Omron HN289 scales. Height was measured to the nearest cm using a Rolson 50565 tape measure with the subject standing against a wall. MAC and humeral length were measured with paper tape measures.

Age-based estimates of weight in kg for children included the original and revised APLS formulae,[1] the finger-counting method,[12] and the Rwanda rule.[7] Broselow, Mercy and PAWPER tape weight estimates were determined ‘virtually’:[13] rather than using the tapes themselves, tables of data for each tape were used to determine appropriate weight estimates in kg for the measured distances in cm. Height was used for four different editions of the Broselow tape: 1993, 1998, 2007, 2011. The Broselow tape extends to approximately 145cm (depending on the version used), and was used only for children. Height and body habitus categories were used for the PAWPER XL tape[11] in children and adolescents; height and MAC for the PAWPER XL-MAC[6] in all subjects. The PAWPER tape extends to 180cm, and subjects up to 199cm are assumed to fall in the same weight category as 180cm. Humeral lengths and MAC were used for the Mercy tape,[4] which was assessed in all subjects. Two MAC formulae were used in all subjects.[3, 9] Finally, Kokong’s height-based formula was used in adults.[10] Table 1 summarises the different methods used in each age-group.

**Table 1.** Weight estimation methods.

| Method | Measurements used | Formula for weight (kg) | Groups included in study |
| --- | --- | --- | --- |
| Original APLS | Age (years) | $(2 \times \text{age}) + 8$ | Children |
| Revised APLS | Age (years) | $(2 \times \text{age}) + 8$ [1 to 6 years old]<br>$(3 \times \text{age}) + 7$ [from 6 years old] | Children |
| Finger-counting | Age (years) | $(2.5 \times \text{age}) + 7.5$ | Children |
| Rwanda rule | Age (years) | $(1.7 \times \text{age}) + 8$ | Children |
| Broselow tape | Height (cm) |  | Children |
| PAWPER XL tape | Height (cm)<br>Body habitus |  | Children<br>Adolescents |
| PAWPER XL-MAC tape | Height (cm)<br>MAC (cm) |  | All |
| Mercy tape | Humeral length (cm)<br>MAC (cm) |  | All |
| 2010 MAC formula | MAC (cm) | $(\text{MAC} - 10) \times 3$ | All |
| 2017 MAC formula | MAC (cm) | $(4 \times \text{MAC}) - 50$ | All |
| Kokong | Height (cm) | Height - 100 | Adults |
| Subject/guardian estimate |  |  | All |

The primary outcome measure for any given method, was the percentage of estimates that lay within 10% (p10), 20% (p20) or 30% (p30) of actual weight.[1, 6, 9] Secondary outcome measures were the Bland-Altman bias (mean percentage error (MPE); a measure of the trueness of the estimate), and limits of agreement (LOA=MPE ± 1.96 SD; a measure of the precision of the estimate).[14] Bland-Altman analysis used the percentage differences between actual and estimated weights.

The p10, p20 and p30 obtained for different methods were compared using the McNemar test for comparison of paired proportions. Bland Altman biases were compared using Wilcoxon’s test for paired samples.

A sample size of 300 for each group was identified as pragmatically realistic for the timescale of the study. Bland Altman analysis requires a sample size of at least 200.[15] For McNemar’s test, 253 subjects are required to detect a difference of 5% in proportions (power 80%, 95% confidence).

MedCalc Statistical Software version 19.1.6 was used for statistical analysis (MedCalc Software Ltd, Ostend, Belgium; https://www.medcalc.org; 2020).

Ethical approval was obtained from the Institutional Review Board, College of Medicine and Health Sciences, University of Rwanda (reference 406/CMHS IRB/2016) and from the Ethics Committee, CHUK (reference EC/CHUK/214/2016). Written consent in Kinyarwanda was obtained from patients or guardians as appropriate.

## RESULTS

947 subjects were recruited: 307 children (1 to 9.9 years), 309 adolescents (10 to 15.9 years) and 331 adults (16.1 to 90 years). All were Rwandan; 442 (46.7%) were female. Age, weight, height and MAC data within each subject group were not normally distributed. Medians and interquartile ranges (IQR) are presented in table 2. Required data were obtained in all subjects, but two children had MAC below, and four adults had humeral length above, the limits of the Mercy tape. Guardians estimated the weight for five of the 16 year old adults.

**Table 2.** Demographic data for each group.

|  | n/N (%) | median | IQR |
| --- | --- | --- | --- |
| **Children** | 307/947 (32%) |  |  |
| Female | 119/307 (39%) |  |  |
| Age (years) |  | 4.5 | 2.6 to 7 |
| Weight (kg) |  | 15.6 | 12 to 21 |
| Height (cm) |  | 100 | 89 to 116 |
| MAC (cm) |  | 15 | 14 to 17 |
| **Adolescents** | 309/947 (33%) |  |  |
| Female | 126/309 (41%) |  |  |
| Age (years) |  | 13.1 | 11.3 to 14.6 |
| Weight (kg) |  | 35 | 28.9 to 43.8 |
| Height (cm) |  | 145 | 133 to 155 |
| MAC (cm) |  | 20 | 18 to 21 |
| **Adults** | 331/947 (35%) |  |  |
| Female | 197/331 (60%) |  |  |
| Age (years) |  | 43.1 | 29.1 to 56.1 |
| Weight (kg) |  | 64 | 56.2 to 75.3 |
| Height (cm) |  | 162 | 157 to 167.8 |
| MAC (cm) |  | 27 | 25 to 30 |

The p10, p20 and p30 for each weight estimation method are presented in table 3. Figure 1 summarises the accuracy of the better weight estimation methods.

**Table 3.** p10, p20 and p30.

|  | ***P10*** |  | ***p20*** |  | ***p30*** |  |
| --- | --- | --- | --- | --- | --- | --- |
|  | ***n*** | ***%*** | ***n*** | ***%*** | ***n*** | ***%*** |
| **Children (n=307)** |  |  |  |  |  |  |
| PAWPER XL | 177 | 57.7 | 252 | 82.1 | 291 | 94.8 |
| PAWPER XL-MAC | 189 | 61.6 | 274 | 89.3 | 297 | 96.7 |
| Broselow 1993 | 174 | 56.7 | 254 | 82.7 | 291 | 94.8 |
| Broselow 1998 | 178 | 58.0 | 265 | 86.3 | 296 | 96.4 |
| Broselow 2007 | 174 | 56.7 | 254 | 82.7 | 291 | 94.8 |
| Broselow 2011 | 166 | 54.1 | 249 | 81.1 | 281 | 91.5 |
| Mercy* | 137 | 44.9 | 230 | 75.4 | 288 | 94.4 |
| 2010 MAC | 97 | 31.6 | 157 | 51.1 | 201 | 65.5 |
| 2017 MAC | 37 | 12.1 | 79 | 25.7 | 126 | 41.0 |
| Original APLS | 137 | 44.6 | 219 | 71.3 | 265 | 86.3 |
| Revised APLS | 93 | 30.3 | 163 | 53.1 | 215 | 70.0 |
| Rwanda rule | 111 | 36.2 | 226 | 73.6 | 278 | 90.6 |
| Finger-counting | 95 | 30.9 | 178 | 58.0 | 227 | 73.9 |
| Guardian estimate | 205 | 66.8 | 277 | 90.2 | 296 | 96.2 |
| **Adolescents (n=309)** |  |  |  |  |  |  |
| PAWPER XL | 116 | 37.5 | 199 | 64.4 | 248 | 80.3 |
| PAWPER XL-MAC | 156 | 50.5 | 282 | 91.3 | 307 | 99.4 |
| Mercy | 138 | 44.7 | 243 | 78.6 | 291 | 94.2 |
| 2010 MAC | 50 | 16.2 | 134 | 43.4 | 233 | 75.4 |
| 2017 MAC | 52 | 16.8 | 121 | 39.2 | 193 | 62.5 |
| Guardian estimate | 209 | 67.6 | 281 | 90.9 | 302 | 97.7 |
| **Adults (n=331)** |  |  |  |  |  |  |
| PAWPER XL-MAC | 140 | 42.3 | 277 | 83.7 | 323 | 97.6 |
| Mercy* | 150 | 45.9 | 267 | 81.7 | 315 | 96.3 |
| 2010 MAC | 54 | 16.3 | 154 | 46.5 | 274 | 82.8 |
| 2017 MAC | 134 | 40.5 | 252 | 76.1 | 312 | 94.3 |
| Kokong | 115 | 34.7 | 223 | 67.4 | 290 | 87.6 |
| Subject estimate** | 287 | 86.7 | 326 | 98.5 | 330 | 99.7 |

\*Mercy tape could not be used in all subjects. For children, $n=305$ . For adults, $n=327$ .
\*\*Guardians estimated weight for five of the 16 year olds.

**Figure 1.**
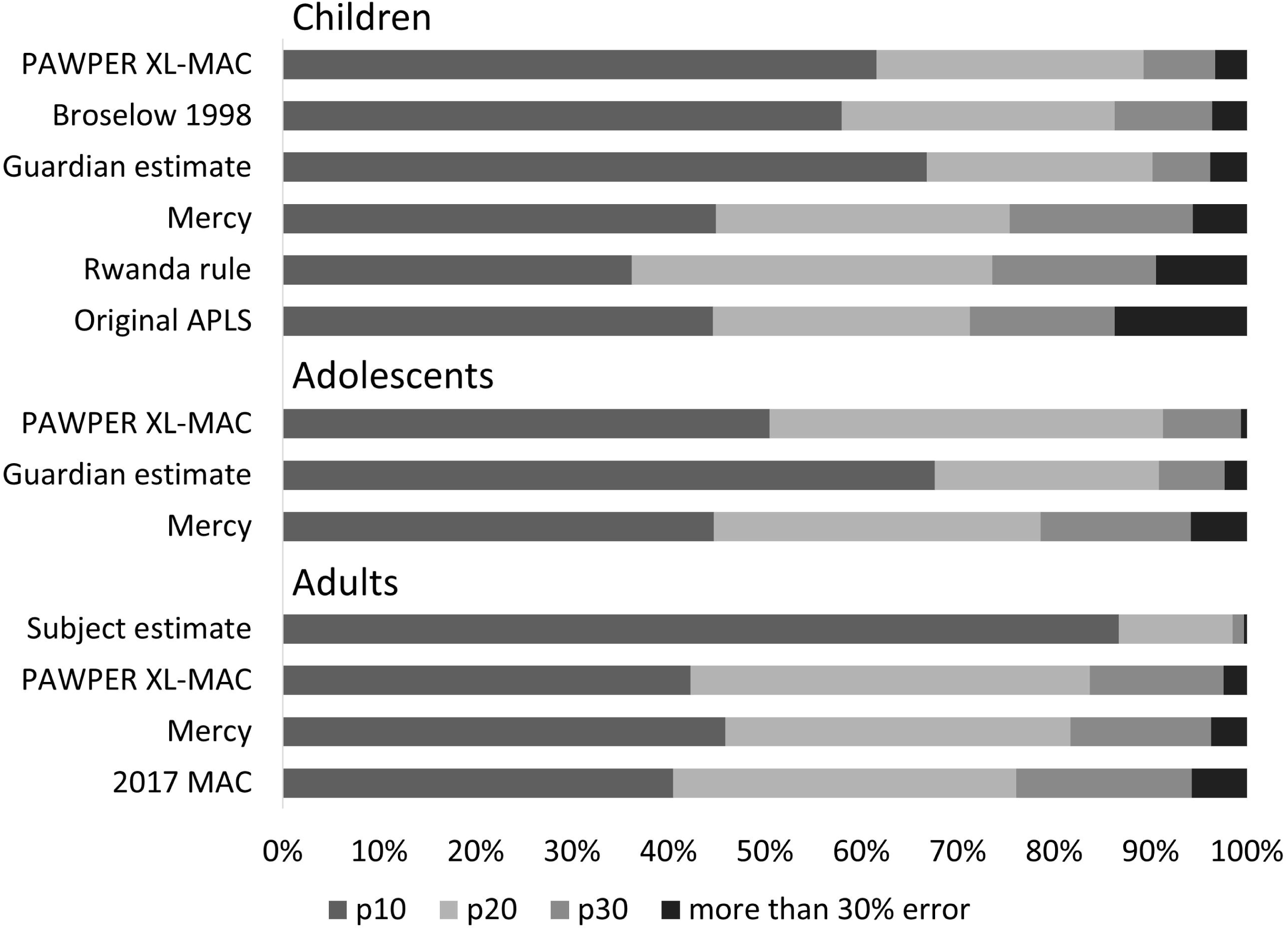
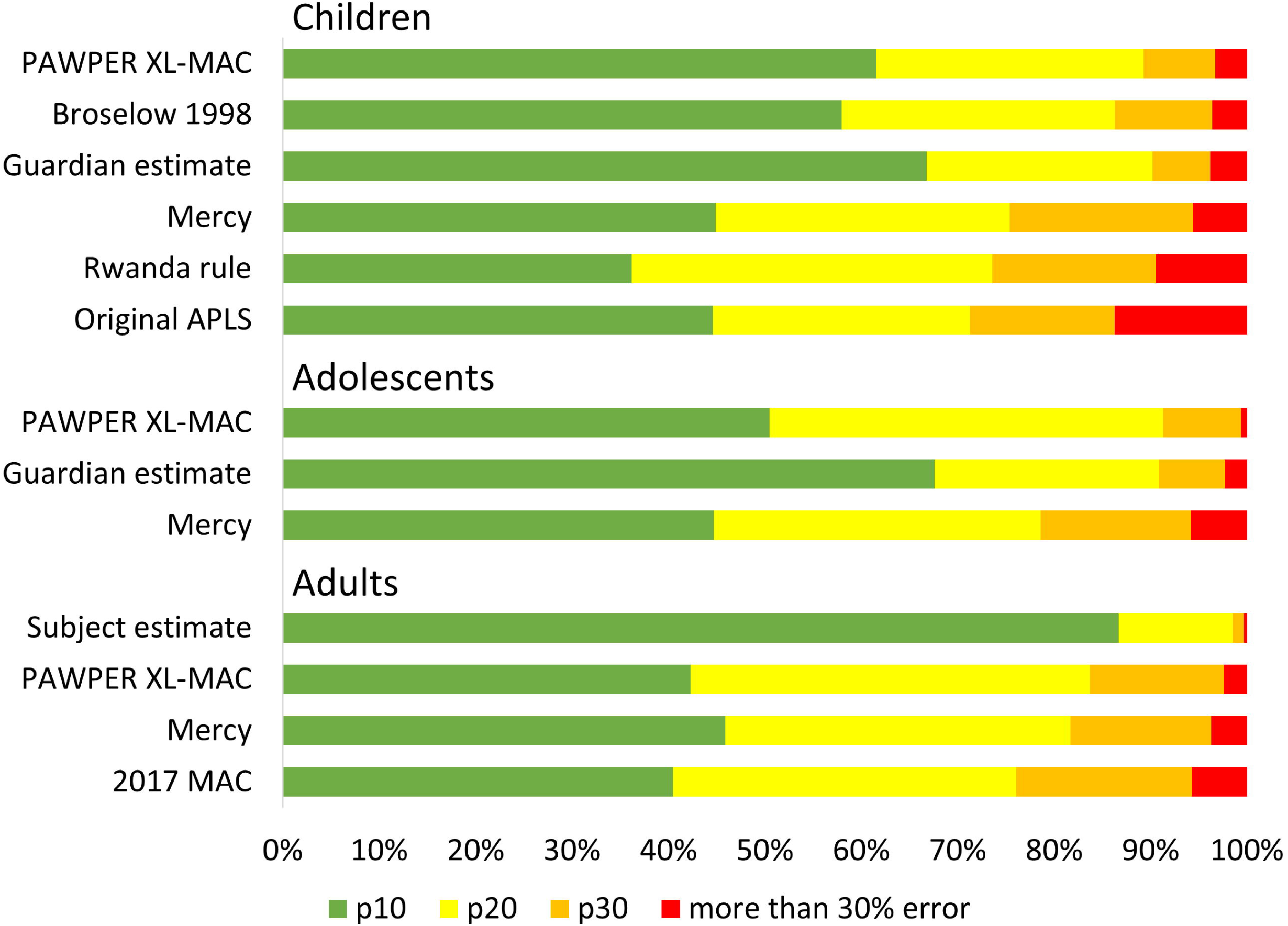
Accuracy of different weight estimation methods. Notes: this figure presents for each method, the proportions of children with weight estimates within 10% (p10), 20% (p20), 30% (p30) of actual weight, and those with an error greater than 30%. For each group of subjects, methods are displayed in descending order of error greater than 30%. Actual percentages are given in table 3.

In children, of the age-based rules, the original APLS formula had the best p10 (44.6% vs Rwanda rule p10=36.2%, p=0.009), and the Rwanda rule had the best p30 (90.6% vs original APLS formula p30=86.3%, p=0.015). There was no difference between their p20. Both were significantly better than the other two age-based formulae, and the two MAC formulae. The Mercy tape had a similar p10 to the original APLS formula (44.9% vs 44.6%), and was not significantly better than the Rwanda rule for p20 (p=0.637) or p30 (p=0.154). Of the different versions of the Broselow tape, the 1998 version performed best (1998 version p20=86.3% vs 1993 version p20=83.7%, p=0.013). This version of the Broselow tape outperformed the Mercy tape (p20=86.3% vs 75.4%, p=0.001), and all age-based and MAC formulae. Even the worst performing Broselow tape (2011 version) outperformed the age-based and MAC formulae. For p20, the PAWPER MAC-XL outperformed the PAWPER XL (89.3% vs 82.1%, p=0.002), and the Mercy tape (89.3% vs 75.4%, p<0.0001). Guardian estimate performed better than the 1998 Broselow tape for p10 (66.8% vs 58%, p=0.021); otherwise there was no difference between that edition of Broselow, the PAWPER XL-MAC and guardian estimate. These were the three best-performing methods in children.

In adolescents, PAWPER XL-MAC outperformed PAWPER XL throughout (for p10, 50.5% vs 37.5%, p=0.002). It outperformed the Mercy tape for p20 (91.3% vs 78.6%, p<0.0001) and p30, but not p10 (p=0.101). Mercy was better than either of the MAC formulae throughout (p<0.0001). Guardian estimate was better than the PAWPER XL-MAC for p10 (67.6% vs 50.5%, p<0.0001), but for p20 and p30 there was no significant difference. In summary, guardian estimate was the best method, followed by the PAWPER XL-MAC and then the Mercy tape.

In adults, the 2017 MAC formula was significantly better than the 2010 formula (p<0.0001 throughout), and was not significantly different from the Mercy tape (for p20, 76.1% vs 81.7%, p=0.057). The Kokong formula was significantly worse than the Mercy tape throughout, and the

2017 MAC formula for p20 and p30. PAWPER XL-MAC was not significantly different from the Mercy tape. Subject estimate was significantly better than either tape (p<0.0001 for p10 and p20; for p30, subject=99.7% vs PAWPER XL-MAC=97.6, p=0.039). In summary, subject estimate was the best method, followed by the PAWPER XL-MAC, Mercy tape, and 2017 MAC formula.

Bland Altman results for the best performing methods are presented in table 4, and an example Bland Altman plot is provided for the PAWPER XL-MAC in children (figure 2). A negative value of the bias represents an under-estimate of actual weight. In children, the Mercy tape bias was significantly larger than any of the other methods (p<0.0001). In adults and adolescents, the guardian/subject estimate bias was significantly smaller than that for the other methods (p<0.0001).

**Table 4.**
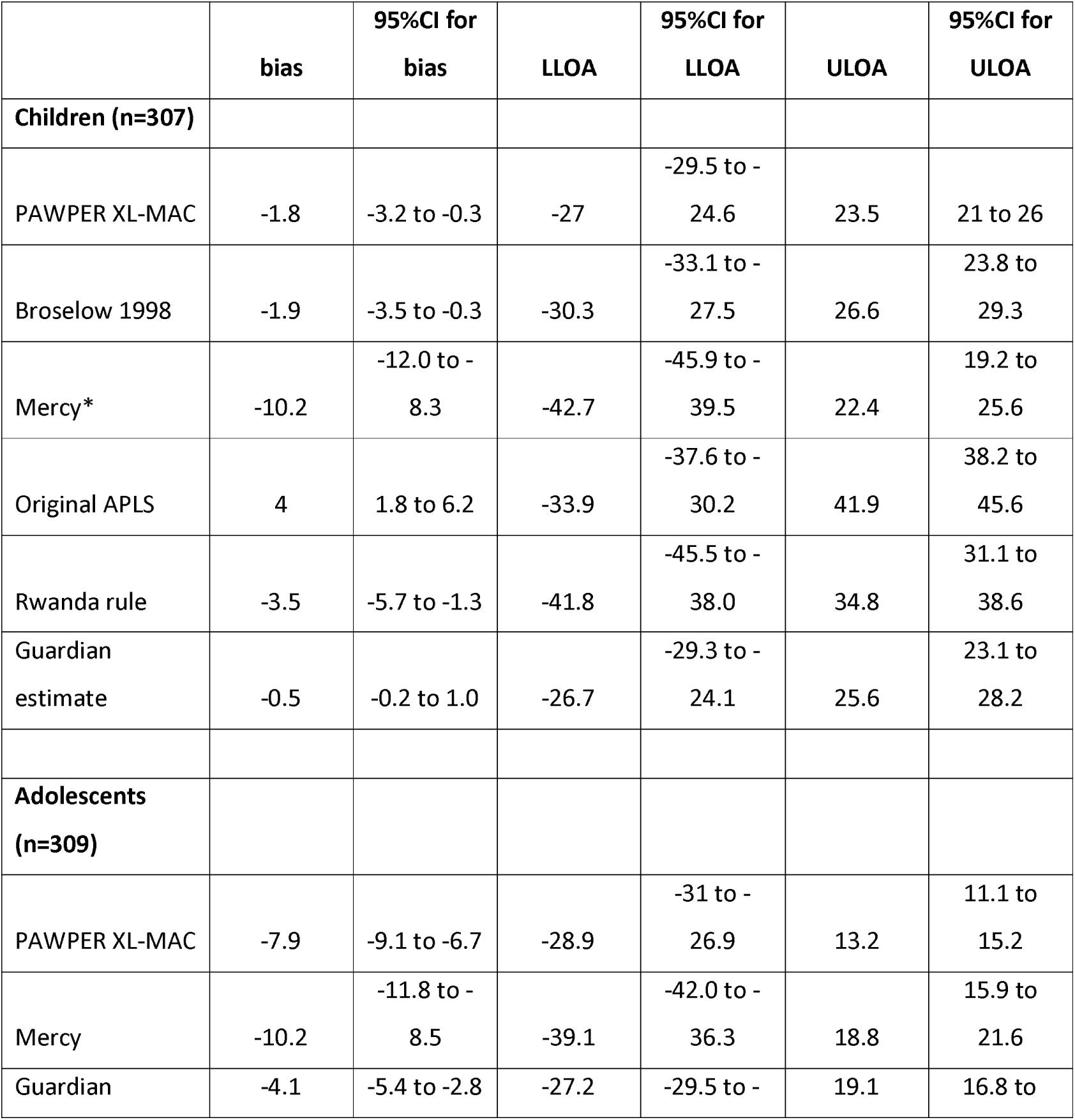
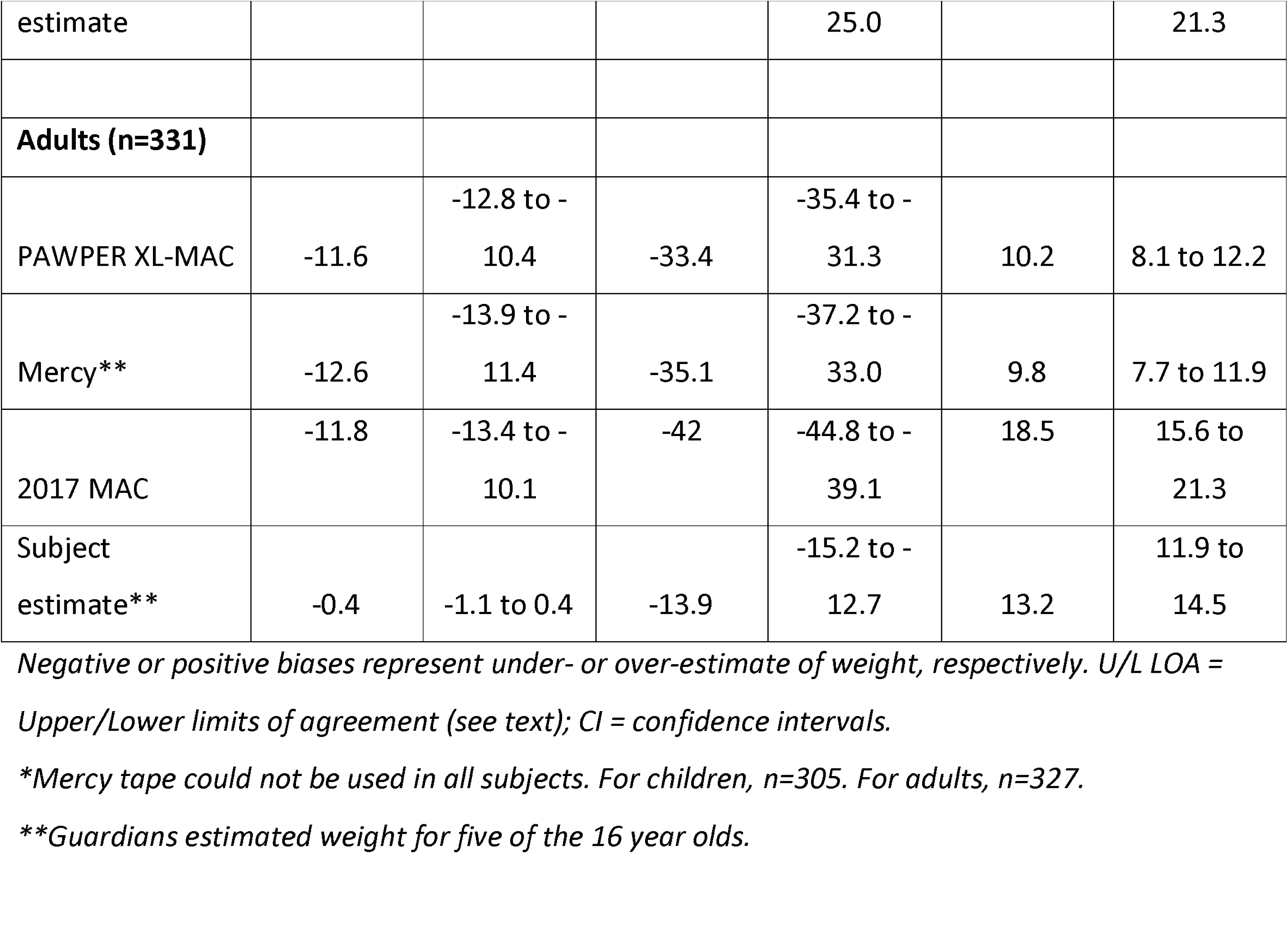
Bland Altman results.

**Figure 2.**
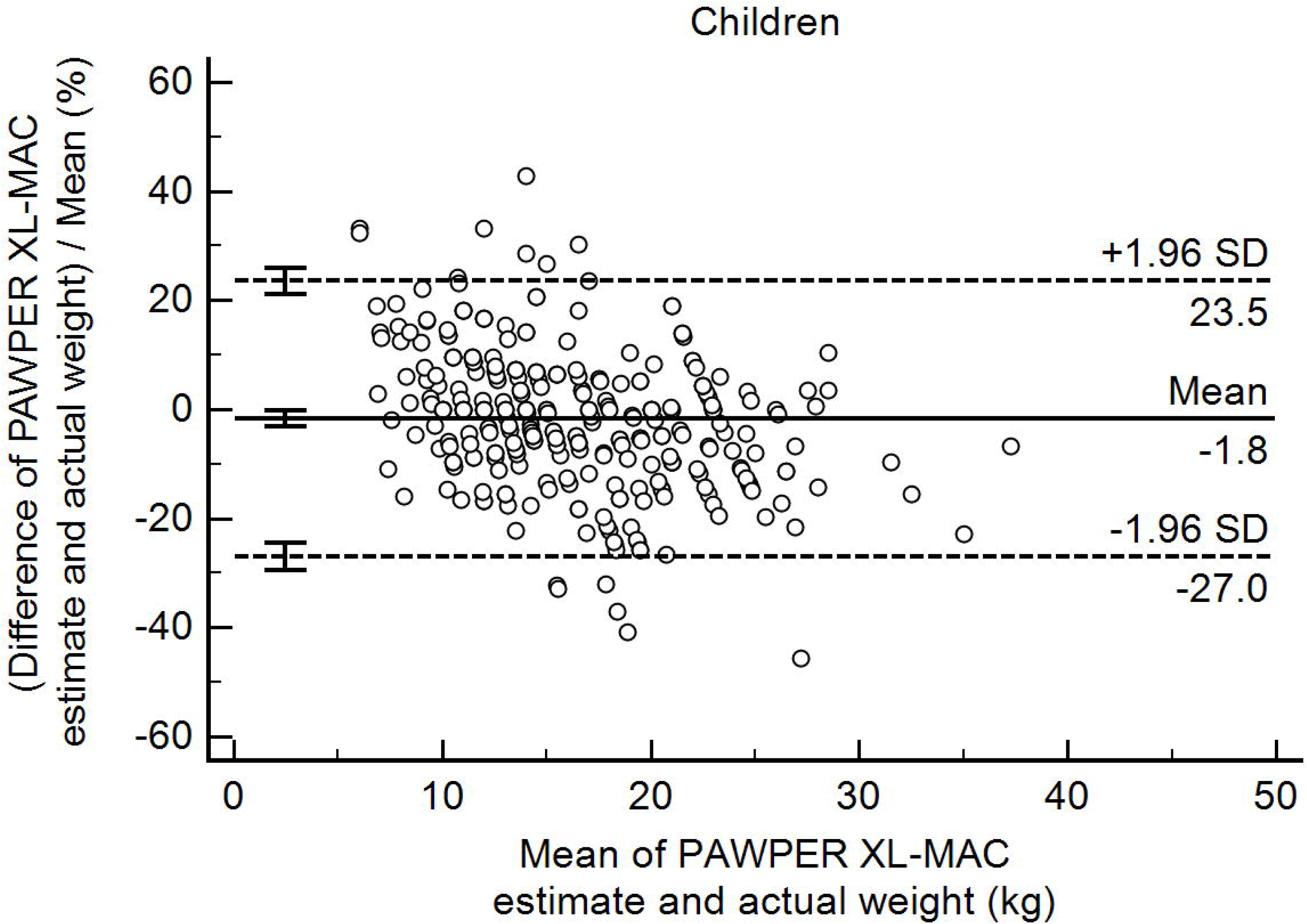
Bland Altman plot for the PAWPER XL-MAC in children. Notes: hatched lines represent LOA. Cross bars depict 95% confidence intervals for LOA and bias. The negative biases represents an average under-estimate of actual weight.

## DISCUSSION

This study has prospectively compared several methods of weight estimation in children, adolescents and adults. The study was well-powered and included large and equally sized age groups. The best methods in children are the PAWPER XL-MAC, the 1998 Broselow tape, and guardian estimate. In adolescents, the best methods are the PAWPER XL-MAC, guardian estimate, and the Mercy tape. In adults, subject estimate is the best method, with the PAWPER XL-MAC, Mercy tape and 2017 MAC formula performing less well. Overall, in terms of the proportion of subjects whose weight estimate is within 10% of actual weight (p10), guardian or subject estimate was significantly better than any other method. We have demonstrated that both the Mercy and PAWPER XL-MAC tapes can be used in adults and adolescents. In children the Rwanda age-based rule has been shown to perform comparably to the best existing alternative, the original APLS formula.

One limitation of the study was that it was conducted in a tertiary referral centre. In the Rwanda healthcare system, patients usually attend this hospital after referral. Some of them may have been weighed before referral, especially the paediatric patients. It is therefore possible that guardian or self-estimation is not as accurate for patients presenting for the first time at district hospitals. A second potential limitation was the gender imbalance, with fewer girls than expected among the children and adolescents, and more women among the adults. This might simply reflect the balance of patients presenting at our hospital. It is unlikely to skew the results significantly, and none of our weight estimation methods includes an option to adjust for gender. Thirdly, the study was virtual, in that we did not use the actual PAWPER, Mercy or Broselow tapes. However, it has been shown that real and virtual studies produce very similar results, although it is recommended to confirm virtual findings in a real-life study.[13]

There is no consensus as to what defines an acceptable degree of accuracy for weight estimation. The original Broselow tape study determined proportions of estimates within 5%, 10%, 15% and 25% but did not argue for one specific cut-off.[2] The MAC formulae papers presented proportions within 10%, 20% and 30%, and suggested that most drugs could safely be given within a margin of error of 30%.[3, 9] Lack argued that as the therapeutic ratio (the toxic dose divided by the effective dose) for most drugs is greater than 1.5, an error in weight-dependent dosage of 10-20% would be reasonable.[16] The creator of the PAWPER tape states that 95% of estimates should lie within 20% of actual weight, and 70% within 10%, for a method to be considered adequate.[5]. Using this standard, none of the methods we assessed would be considered adequate in any age group, other than subject estimation in adults. Other studies consistently show that age-based rules for children would fail that standard, not even reaching a p30 of 90%.[1, 5] The original Broselow tape study only found a p10 of 59.7%. If we were to define acceptability as a p30 of 95% (ie, that 95% of estimates lie within 30% of actual weight), then in Rwandan children we would recommend only the PAWPER XL-MAC, the 1998 Broselow tape, and guardian estimation. In adolescents we would recommend only the PAWPER XL-MAC and guardian estimation. In adults we would recommend the Mercy tape, PAWPER XL-MAC and subject estimation. By any of these definitions of adequacy, none of the age-based rules would be acceptable in children, nor would Kokong’s height-based formula in adults, nor would either of the MAC formulae. If there were no guardian available, and no tape, then we would recommend cautious use of the original APLS formula in children. Although it did not significantly outperform the new Rwanda rule overall, it is much easier to calculate.

A recent study has found that the MAC formula did not perform as well in an elderly Dutch population as it did in an American population, highlighting the need for methods based on local population data.[17] However, ‘1-dimensional’ (1D) methods that use only one body-based measure (eg, height or MAC) are consistently outperformed by ‘2-dimensional’ (2D) methods that utilise two different body measurements: a length-based measure, and a habitus measure.[6] There are two 2D methods in use, the Mercy and PAWPER tapes. These methods are inherently more precise: for any given height, people could have a wide range of weights. But for any given height together with a given MAC, the range of weights is much narrower. Previous Mercy and PAWPER studies have demonstrated high values of p10 and p20 (78.6% and 98% for Mercy, 79.3% and 96.9% for PAWPER XL-MAC).[4, 6] In our study, Bland Altman analysis confirms that the PAWPER MAC-XL has narrower LOA in children than Broselow, MAC or age-based methods. However, we have not obtained as good overall accuracy (p10, p20) in our study as in the original PAWPER and Mercy studies. This is likely due to population differences, and reinforces the need for local studies. Encouragingly, the PAWPER XL-MAC has narrow LOA in adolescents and adults, which suggests that adapting the tape to improve the bias (based on population median heights and MAC), should improve overall accuracy in these ages.

Wells has noted that the only method as accurate as 2D methods, is parental estimate.[6] We have confirmed that this is true in children and adolescents, and that subject estimate is the best in adults too. However, in emergency situations patients might not be able to estimate their weight and guardians may not be present, which is why alternative methods of weight estimation are essential. This study has not addressed whether clinician estimate is reliable in our context. Previously, clinician estimate has been found to be significantly worse than patient or parent estimates in adults or children.[18, 19] For now, it appears that the PAWPER XL-MAC is the overall best method to use in all ages.

## Conclusions

Both the PAWPER and Mercy tapes can be used in adolescents and adults. In children, the new Rwanda age-based rule performed comparably to the original APLS formula, but neither were considered acceptable. If no other option is available, we would recommend cautious use of the APLS formula. Across all ages, the PAWPER XL-MAC and guardian/subject estimates of weight were the most reliable and we would recommend their use in this setting, with the proviso that the PAWPER XL-MAC (and Mercy) tape might benefit from adjustment for local population data, with subsequent real-life validation.

## Data Availability

Deidentified Excel/Medcalc datasets are available on request from the author (ORCID ID 0000-0002-8910-2307).

## Dissemination of results

These results were presented as a poster at the Royal College of Emergency Medicine annual scientific conference, October 2017.

## Authors’ contributions

Both authors were involved in the planning of the study, and the ethics and grant application processes. AM supervised data collection and reviewed the manuscript. GC analysed the data and wrote the manuscript. Both authors approved the final version of the manuscript.

